# First “snap-shot” meta-analysis to estimate the prevalence of serum antibodies to SARS-CoV-2 in humans

**DOI:** 10.1101/2020.08.31.20185017

**Authors:** Ali Rostami, Mahdi Sepidarkish, Mariska Leeflang, Seyed Mohammad Riahi, Malihe Nourollahpour Shiadeh, Sahar Esfandyari, Ali H Mokdad, Peter J. Hotez, Robin B. Gasser

**Author notes:** **Correspondence to:** Assistant Professor Ali Rostami, Infectious Diseases and Tropical Medicine Research Center, Health Research Institute, Babol University of Medical Sciences, Babol, Iran. Professor Robin B. Gasser, Department of Veterinary Biosciences, Melbourne Veterinary School, The University of Melbourne, Parkville, Victoria, Australia.

## Abstract

**Background:** COVID-19 is arguably the number-one public health concern worldwide, and efforts are now escalating to control its spread.

**Objective:** In this study, we undertake a meta-analysis to estimate the global and regional anti-SARS-CoV-2 seroprevalence rates in humans and assess whether seroprevalence associates with geographical, climatic and socio-demographic factors.

**Data sources:** We systematically reviewed PubMed, Scopus, Embase, medRxiv and bioRxiv for peer-reviewed articles or preprints (up to 14 August 2020).

**Study eligibility criteria:** Population-based studies describing prevalence of anti-SARS-CoV-2 serum antibodies in general people.

**Participants:** general people who were tested for prevalence of anti-SARS-CoV-2 serum antibodies.

**Interventions:** There were no interventions.

**Methods:** We used random-effects model to estimate pooled seroprevalence, and then extrapolated these findings to the global population (for 2020). Sub-group and meta-regression analyses explored potential sources of heterogeneity in the data and relationships between seroprevalence and socio-demographic, geographical and climatic factors.

**Results:** In total, 47 serological studies involving 399,265 people from 23 countries met the inclusion criteria. The pooled seroprevalence of SARS-CoV-2 in general people was estimated at 3.38% (95% CI, 3.05%–3.72%; 15,879/399,265). On a regional basis, we determined seroprevalence estimates of 5.27% (3.97–6.57%) in Northern Europe; 4.41% (2.20–6.61%) in Southern Europe; 4.41% (3.03– 5.79%) in North America; 3.17% (1.96–4.38%) in Western Europe; 2.02% (1.56–2.49%) in the Eastern Asia; and 1.45% (0.95–1.94%) in South America. Extrapolating to the 2020 world population, we estimated that 263,565,606 individuals had been exposed or infected with SARS-CoV-2 at the first wave of the pandemic. A significantly higher seroprevalence was related to higher income levels and human development indices, higher geographical latitudes and lower mean environmental temperatures.

**Interpretation:** This study reinforces that SARS-CoV-2 infection is a very rapidly-spreading communicable disease and calls for routine surveys to constantly monitor temporal changes in seroprevalence around the globe.

## Introduction

COVID-19, a severe, acute respiratory syndrome caused by the coronavirus SARS-CoV-2, was identified, for first time, in December, 2019, in Wuhan, China [1, 2] and, within months, spread to most nations of the world [3]. By 16 August 2020, this pandemic disease was affecting people in 213 countries and territories, with ~ 21 million laboratory-confirmed cases and ~ 800,000 deaths reported globally [4]. The diagnosis and management of COVID-19 is based on the detection of SARS-CoV-2 in nasopharyngeal swabs from patients presenting with clinical signs (including fever, dry cough and/or shortness of breath), or in suspected persons, by reverse transcription-polymerase chain reaction (RT–PCR) [5, 6]. Since the manifestation of SARS-CoV-2 infection ranges from asymptomatic to fatal, epidemiological surveillance of confirmed COVID-19 cases might not be representative for a particular community [7, 8]. Although RT-PCR is presently considered a “gold standard” for the diagnosis of SARS-CoV-2 infection [5], a significant number of subclinical and asymptomatic, infected individuals are likely to remain undetected. Therefore, it is plausible or likely that the actual number of people exposed to, or infected with, is underestimated [7-9]. Serological screening represents a critical adjunct to PCR-based detection/diagnosis and is a key tool to evaluate the cumulative prevalence of SARS-CoV-2 infection, and to monitor seroconversion [10] and seroreversion [11, 12] in individuals and a community, in order to gain knowledge and understanding of the dynamics of specific antibody responses during and after the spread of the virus and, if undertaken routinely, to inform health authorities, politicians and policy makers about seroprevalence at any given stage during an epidemic [13, 14]. The prevalence of specific serum antibodies (IgG or IgM) against SARS-CoV-2 can provide a sound indication of exposure to SARS-CoV-2 in a population [7, 9]. Due to the persistence of antibodies to SARS-CoV-2 (in particular IgG) after viral clearance [7], it is expected that serological monitoring and surveillance are providing highly relevant data sets to estimate the cumulative prevalence of SARS-CoV-2 infection/exposure in a population [7, 15], and may even indicate the immune status of individuals or populations [8, 9].

Several commercial and in-house immunoassays are being used for the detection of IgM and/or IgG antibodies to SARS-CoV-2; these are mainly enzyme-linked immunosorbent assays (ELISAs), chemiluminescence immunoassays (CLIAs) or lateral flow assays (LFIAs) [16, 17]. The diagnostic sensitivity and specificity of these methods vary, and depend on the use of recombinant or purified protein antigen (e.g., spike (S), envelope (E), membrane (M), nucleocapsid (N) or receptor binding domain (RBD) proteins), and the rigor of assay optimization [18, 19].

In past weeks, sero-epidemiological studies have been reported from a number of countries most affected by COVID-19, including Brazil, China, France, Germany, Iran, Italy, Spain, England and the USA [9, 20-27]. Although the pandemic is still spreading, it is crucial that a rapid and thorough analysis be undertaken to estimate global seroprevalence at moment in times. In this study, six months after the commencement of the pandemic, we undertake a meta-analysis to estimate the global and regional seroprevalences of SARS-CoV-2 in the general population, and assess whether geographical, climatic and socio-demographic factors impact on seroprevalence.

## Methods

### Search strategy and selection criteria

This study followed the Preferred Reporting Items for Systematic Reviews and Meta-Analyses (PRISMA) guidelines (cf. Figure 1). We performed a systematic literature search in PubMed, Scopus, Embase, medRxiv and bioRxiv on August, 2020 using the following terms: “SARS-CoV-2”, “COVID-19”, “coronavirus”, “antibody”, “ELISA”, “seroprevalence”, and “population”. Additional related articles were retrieved from Google Scholar and manual review of included papers. All articles were imported to Endnote software X8 (Thompson and Reuters, Philadelphia, PA), and duplicates were removed. Two independent reviewers (A.R., M.S.) screened all titles and abstracts for eligibility. Included were all peer-reviewed population-based studies, preprints, and research reports (published in English) which reported the prevalence of anti-SARS-CoV-2 serum antibodies in a ‘general’ population. We excluded articles if they (1) recruited suspected, confirmed or hospitalised patients; (2) were performed in at-risk population (e.g., health-care workers) or individuals with known diseases (e.g., cancer or dialysis patients); (3) recorded prevalence based on clinical manifestation, computed tomography scan or PCR; (4) were comparative studies of diagnostic methods; (5) were case reports or case studies; (6) were editorials, commentaries, reviews, systematic reviews without original data.

**Figure 1.**
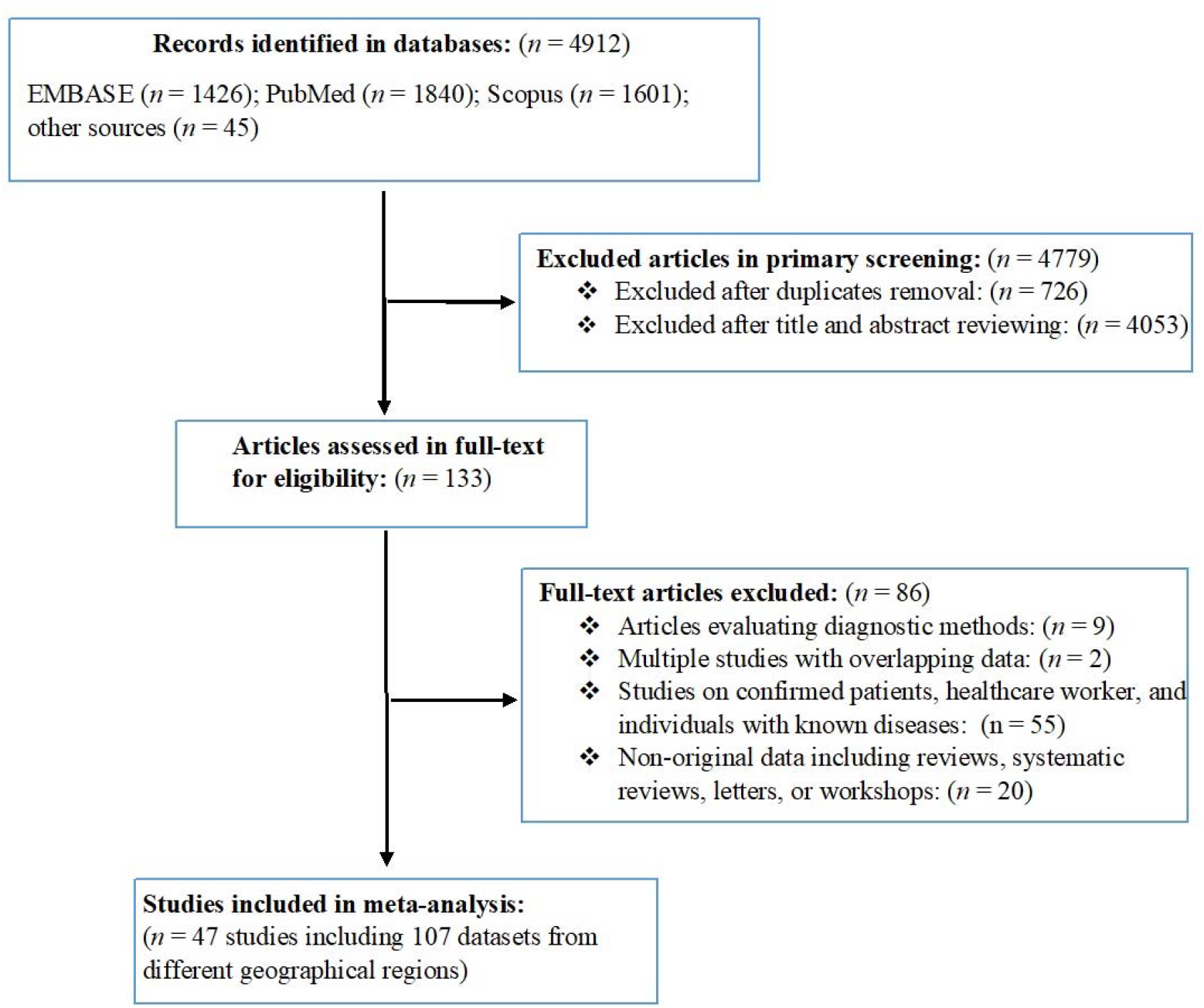
Flowchart of the search strategy and study-selection process, indicating numbers of studies (and associated datasets) excluded or included.

### Data extraction and quality assessment

After the screening of published articles for eligibility, a specific form in Microsoft Excel (version 2016; Microsoft Corporation, Redmond, WA) was used to extract relevant data and information from each eligible study. Two authors (A.R. and M.N.S) independently extracted the required data from eligible studies, and two others (M.S. and S.E.) independently verified these data. We resolved inconsistencies by consensus. The following items were extracted from each study (if described): first author; year of publication; country; city; study period; study design; type of serological methods used; sensitivity and specificity of diagnostic methods; number of people screened; number of people seropositive for SARS-CoV-2 antibodies and data regarding to age, sex and race.

All geographical areas (i.e. countries and cities) studied were classified according to ‘Sustainable Development Goal’ (SDG)-regions or sub-regions defined by the United Nations [28]. For each country, we also recorded information on the total numbers of confirmed cases and deaths (up to 15 August 2020) reported by World Health Organization (WHO) [29], World Bank’s income category [30], gross national income per capita [31] and the human development index [HDI] [32]. Furthermore we recorded total global, regional and national populations (both sexes combined) in 2020 estimated by the United Nations [33]. If sample size(s) and the numbers of sero-positive people were specified in studies, we extracted and appraised data for separate regions. We also recorded information on latitude, longitude, mean relative humidity, and mean environmental temperature in geographic regions/subregions during the study period using the database timeanddate.com (weblink: https://www.timeanddate.com). The quality of studies included in the meta-analysis was assessed using the Joanna Briggs Institute (JBI) Prevalence Critical Appraisal Tool.[34] Individual articles were assessed as to whether they adequately described: sample collection, recruitment method, study subjects and the setting, number of subjects, information on subjects, results, reliability of results, statistical analysis method(s), subpopulation analysis and confounder adjustment (“yes” or “no” answer). For each study, the number of “yes” answers to these 10 criteria was counted; the higher the number of “yes” answers, the lower the risk of bias in a study.

### Meta-analysis

All analyses were performed using Stata statistical software (v.13 Stata Corp., College Station, TX, USA). To conservatively estimate the pooled seroprevalence of SARS-CoV-2 in general people, we used a DerSimonian and Laird random-effects models (REM) [35]. For this purpose, first, we estimated the seroprevalence in individual countries by synthesizing the seroprevalence rates of all studies from the same country, and then calculated the seroprevalences of SARS-CoV-2 for the WHO-defined-regions (if studies were available for at least two countries) by synthesizing the data across individual countries within the same WHO-defined-region. We calculated the pooled seroprevalence rates at a 95% confidence interval (CI) using metaprop command in Stata software. We estimated the amount of heterogeneity using the *I*^2^ statistic, and an *I*^2^ of >75% and a p < 0.05 was considered substantial heterogeneity [36]. To obtain the number of people exposed to SARS-CoV-2, we extrapolated our prevalence estimates to the total human population (in 2020) living in countries and regions – according to the UN Population Division [28].

To explore the potential sources of heterogeneity and also effects of socio-demographic, geographical and climatic parameters on seroprevalences of SARS-CoV-2, we undertook several sub-group analyses by REM and as well as random effects meta-regression ecological analyses using the metareg STATA command [37]. These analyses were performed considering SDG-regions, type of serological method used, age, sex, race, country income level, country HDI, latitude, longitude, mean temperature and/or mean relative humidity. To assess the effect of these variables on seroprevalence, we performed random effects meta-regression analyses using the metareg STATA command [37]. Further meta-regression analyses were performed to see whether seroprevalences rate is associated with number of total confirmed or total deaths in countries. Results were statistically significant if the *P* value was < 0.1.

## Results

### Study characteristics

A search of electronic databases yielded a total of 4,912 articles; following the removal of duplicate articles and a review of article titles and abstracts, 133 potentially relevant articles were identified for full-text evaluation (Figure 1). After applying the eligibility criteria, 47 articles met the selection criteria to be included in the quantitative synthesis. These 47 eligible articles contained 107 datasets representing 399,265 people from 23 countries in six SDG-regions. Of these datasets, 74 were from Europe and Northern America, 17 from Latin America and the Caribbean, 13 from Eastern and South-eastern Asia, one from Central and Southern Asia, one from North Africa and Western Asia, and one from Sub-Saharan Africa. We did not identify a published study from Oceania. The characteristics of studies included are given in Supplementary Table 1. The majority of the articles included (44 studies) had a low risk of bias (score: 7-10/10), and only three studies had moderate risk (6/10) of bias (Supplementary Table 2).

### Seroprevalence of SARS-CoV-2

This analysis of the 107 datasets selected from the 47 articles showed that 15,879 people from a general population of 399,265 had specific serum antibodies to SARS-CoV-2, indicating a pooled seroprevalence of 3.38% (95% CI, 3.05%–3.72%). The heterogeneity among studies was substantial (*I*^2^ = 99.4%, *P* < 0.001. After extrapolation to the 2020 world population, we estimated that 263,565,606 (237,741,369–289,966,523) individuals were exposed to SARS-CoV-2 (13 July 2020). More detail on the overall and regional seroprevalences and burden of SARS-CoV-2 seropositivity to SARS-CoV-2 is given in Table 1. According to SDG-subregions (for which ≥ 2 countries were represented), seroprevalences were 5.27% (3.97–6.57%) in Northern Europe; 4.41% (2.20–6.61%) in Southern Europe; 4.41% (3.03–5.79%) in Northern America; 3.17% (1.96–4.38%) in Western Europe; 2.02% (1.56–2.49%) in Eastern Asia; and 1.45% (0.95–1.94%) in South America. Countries with the highest seroprevalences were Iran (22.1%), Sweden (15.02), Chile (10.7%), Switzerland (7.9%), Italy (7.27%), South-Korea (7.5%), Spain (5.0%) and USA (4.4%). Figure 2 shows a geographic information system (GIS) map summarising the seroprevalence estimates of SARS-CoV-2 in the general population in individual countries.

**Table 1.** Global, regional and national pooled prevalence of serum antibodies to SARS-CoV-2 in the general population (results from 47 studies containing 107 datasets performed in 23 countries).

| WHO regions/<br>country | Number<br>datasets | Number of<br>people screened<br>(total) | Number of<br>seropositive<br>people | Pooled<br>seroprevalence<br>% (95% CI) | Estimated global<br>or country's<br>population (2020) | Estimated number of people exposed<br>to SARS-CoV-2 |
| --- | --- | --- | --- | --- | --- | --- |
| **Global** | **107** | **399,265** | **15,879** | **3.38 (3.05–3.72)** | **7,794,799,000** | **263,565,606 (237,741,369–289,966,523)** |
| **Europe and**<br>**northern America** | **74** | **272,265** | **13,109** | **4.21 (3.52–4.90)** | **1,116,506,000** | **47,004,902 (39,301,011–54,708,794)** |
| **Northern America** | **22** | **51,544** | **3,146** | **4.41 (3.03–5.79)** | **368,870,000** | **16,267,167 (11,176,761–21,357,573)** |
| United states | 22 | 51,544 | 3,146 | 4.41 (3.03–5.79) | 331,003,000 | 14,597,232 (10,029,390–19,165,073) |
| **Western Europe** | **13** | **16,933** | **658** | **3.17 (1.96–4.38)** | **196,146,000** | **6,217,828 (3,844,461–8,591,194)** |
| Belgium | 2 | 7,391 | 293 | 3.46 (3.04–3.88) | 11,590,000 | 401,014 (352,336–449,692) |
| France | 5 | 1,198 | 30 | 2.19 (1.20–3.18) | 65,274,000 | 1,429,500 (783,288–2,075,713) |
| Germany | 4 | 3,806 | 81 | 2.23 (0.79–3.67) | 83,784,000 | 1,868,388 (661,893–3,074,872) |
| Switzerland | 1 | 2,766 | 219 | 7.92 (6.94–8.99) | 8,655,000 | 685,476 (860,307–) |
| Luxembourg | 1 | 1,862 | 35 | 1.88 (1.31–2.60) | 626,000 | 11,768,000 (8,200–16,276) |
| **Southern Europe** | **26** | **71,478** | **3,242** | **4.41 (2.20–6.61)** | **152,215,000** | **6,712,681 (3,348,730–10,061,411)** |
| Croatia | 2 | 1,494 | 19 | 1.05 (0.56–1.60) | 4,105,000 | 43,102 (22,988–65,680) |
| Italy | 4 | 2,323 | 145 | 7.27 (2.48–11.9) | 60,462,000 | 4,395,587 (1,499,457–7,249,393) |
| Spain | 19 | 61,075 | 3,054 | 5.01 (4.83–5.18) | 46,755,000 | 2,342,425 (2,258,266–2,421,909) |
| Greece | 1 | 6,586 | 24 | 0.36 (0.23–0.54) | 10,423,000 | 37,522 (23,972–56,284) |
| **Eastern Europe** | **1** | **10,474** | **69** | **0.66 (0.51–0.83)** | **293,013,000** | **1,933,885 (1,494,366–2,432,007)** |
| Hungary | 1 | 10,474 | 69 | 0.66 (0.51–0.83) | 9,660,000 | 63,756 (49,266–80,178) |
| **Northern Europe** | **12** | **121,836** | **5,994** | **5.27 (3.97–6.57)** | **106,261,000** | **5,599,954 (4,218,561–6,981,347)** |
| England | 9 | 99,908 | 5,544 | 5.65 (4.61–6.69) | 67,886,000 | 3,835,559 (3,129,544–4,541,573) |
| Denmark | 2 | 21,715 | 418 | 1.77 (1.60–1.95) | 5,792,000 | 102,518 (92,672–112,944) |
| Sweden | 1 | 213 | 32 | 15.0 (10.5–20.5) | 10,099,000 | 1,516,869 (1,061,405–2,074,334) |
| **Eastern and**<br>**south-eastern Asia** | **13** | **89,648** | **1,855** | **2.02 (1.56–2.49)** | **2,346,709,000** | **47,403,521 (36,608,660– 58,433,054)** |
| **Eastern Asia** | **12** | **88832** | **1852** | **2.02 (1.56–2.49)** | **1,678,090,000** | **33,897,418 (26,178,204– 41,784,441)** |
| China | 8 | 86,416 | 1,756 | 1.63 (1.13–2.13) | 1,439,324,000 | 23,460,981 (16,264,361 – 30,657,601) |
| Japan | 3 | 2,218 | 81 | 3.62 (2.84–4.39) | 126,476,000 | 4,578,431 (3,591,918 – 5,552,296) |
| South-Korea | 1 | 198 | 15 | 7.58, (4.30–12.2) | 51,269,000 | 3,886,190 (2,204,567 – 6,249,691) |
| **South-Eastern**<br>**Asia** | **1** | **816** | **3** | **0.37 (0.08–1.07)** | **668,620,000** | **2,473,894 (534,896–7,154,234)** |
| Malaysia | 1 | 816 | 3 | 0.37 (0.08–1.07) | 32,366,000 | 119,754 (25,893–346,316) |
| **Latin America**<br>**and the Caribbean** | **17** | **33,596** | **618** | **1.45 (0.95–1.94)** | **653,962,000** | **9,482,449 (6,212,639–12,686,862)** |
| **South America** | **17** | **33,596** | **618** | **1.45 (0.95–1.94)** | **430,760,000** | **6,246,020 (4,092,220–8,356,744)** |
| Brazil | 15 | 32,352 | 479 | 0.96 (0.52–1.40) | 212,559,000 | 2,040,566(1,105,306– 2,975,826) |
| Chile | 2 | 1,244 | 139 | 10.78 (9.1–12.5) | 19,116,000 | 2,060,704(1,731,909 –2,389,500) |
| **Sub-Saharan**<br>**Africa** | **1** | **3,098** | **174** | **5.62 (4.83–6.49)** | **1,094,366,000** | **61,503,369 (52,857,878–71,024,353)** |
| Kenya | 1 | 3,098 | 174 | 5.62 (4.83–6.49) | 53,771,000 | 3,021,930 (2,597,139–3,489,738) |
| **Central and**<br>**southern Asia** | **1** | **528** | **117** | **22.16 (18.7–26.0)** | **2,014,709,000** | **446,459,514(376,549,112– 522,816,985)** |
| Iran | 1 | 528 | 117 | 22.16 (18.7–26.0) | 83,993,000 | 18,612,848(15,698,291–21,796,183) |
| **Northern Africa**<br>**and western Asia** | **1** | **130** | **6** | **4.62 (1.71–9.78)** | **525,869,000** | **24,295,147(8,992,359– 51,429,988)** |
| Libya | 1 | 130 | 6 | 4.62 (1.71–9.78) | 6,871,000 | 317,440 (117,494 – 671,983) |

**Figure 2.**
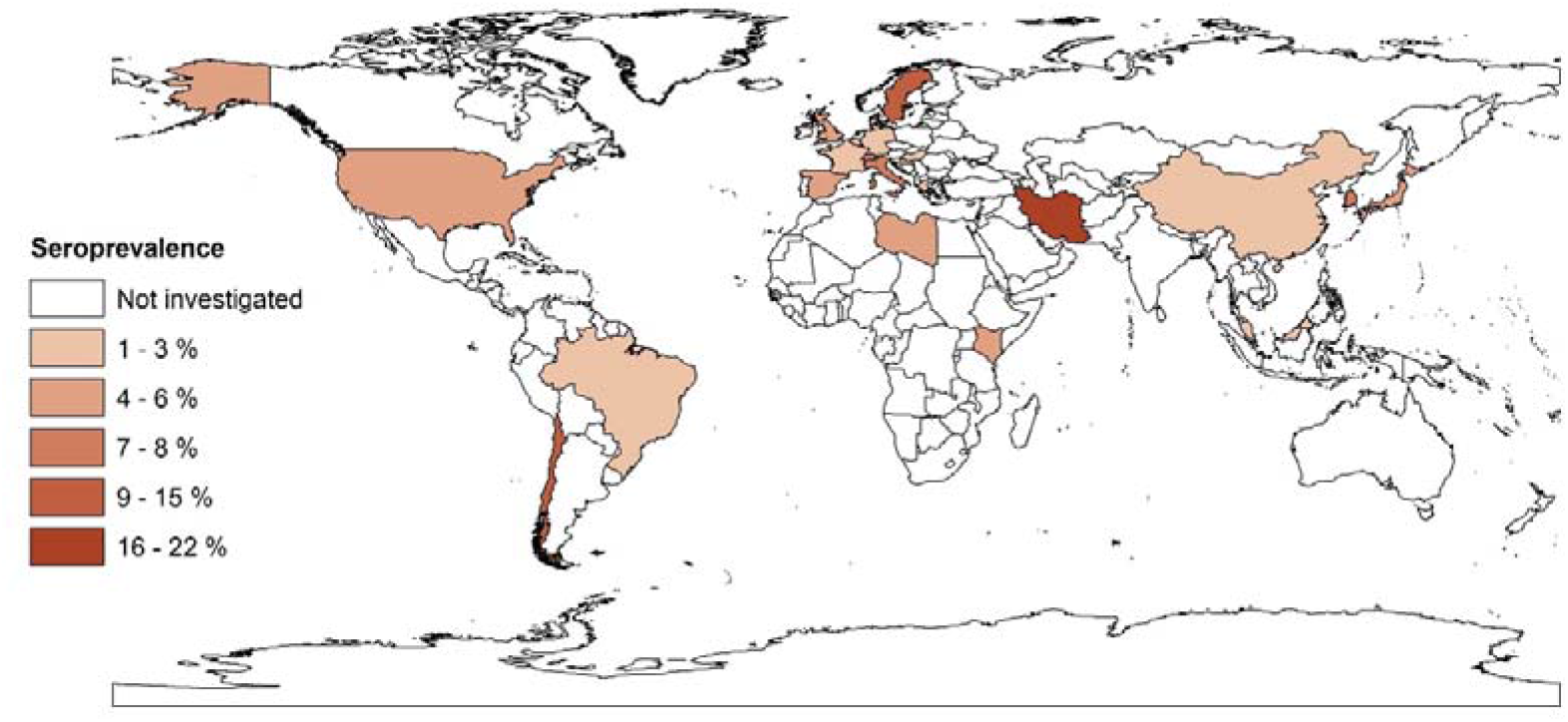
Estimated prevalences of anti-SARS-CoV-2 serum antibodies in the general human population in different countries using geographic information system (GIS).

### Seroprevalence according to sex, age and population

Of the 47 studies included, 29 reported separate, pooled seroprevalences for males and females. Of 145,368 males and 151,790 females, 6,186 males (5.33%, 4.35–6.31%) and 6,958 females (5.05%, 4.06–6.04) had specific serum antibodies against SARS-CoV-2. Fifteen studies reported pooled seroprevalences for different age groups; subgroup analyses revealed pooled seroprevalences in the general population of 2.28% (1.01–3.56%), 3.22% (1.90–4.55%), 2.98% (1.59–4.36%) and 2.57% (1.39–3.76%) in people of ≤19, 20-49, 50-64 and ≥ 65 years of age, respectively (Table 2).

**Table 2.** Prevalence of serum antibodies to SARS-CoV-2 in the general population according to *a priori* defined subgroups

| Variable/subgroups | Number datasets | Number of people screened (total) | Number of seropositive people | Pooled seroprevalence % (95% CI) |
| --- | --- | --- | --- | --- |
| **Gender** |  |  |  |  |
| Male | 29 | 145,368 | 6,186 | 5.33 (4.35–6.31) |
| Female | 29 | 151,790 | 6,958 | 5.05 (4.06–6.04) |
| **Age** |  |  |  |  |
| ≤19 | 11 | 18,333 | 535 | 2.28 (1.01–3.56) |
| 20–49 | 15 | 96,109 | 4,268 | 3.22 (1.90–4.55) |
| 50–64 | 15 | 75,589 | 3,769 | 2.98 (1.59–4.36) |
| ≥65 | 12 | 41,421 | 1,634 | 2.57 (1.39–3.76) |
| **Type of population** |  |  |  |  |
| General | 68 | 227,428 | 6,483 | 2.43 (2.16–2.70) |
| General-adult | 18 | 169,016 | 9,201 | 5.31 (4.12–6.50) |
| General-children | 2 | 1,821 | 162 | 8.76 (7.46–10.06) |
| **Serological method** |  |  |  |  |
| LFIA | 58 | 224,922 | 10,023 | 3.95 (3.17–4.74) |
| ELISA | 23 | 38,159 | 1,417 | 3.53 (2.65–4.40) |
| CLIA | 15 | 80,435 | 1,907 | 2.73 (2.03–3.42) |
| Virus neutralisation assay | 10 | 40,648 | 645 | 1.32 (0.90–1.74) |
| Microsphere immunoassay | 1 | 15,101 | 1,887 | 12.50 (11.97–13.03) |
| **Type of procedure** |  |  |  |  |
| Commercial kit | 83 | 334,334 | 13,870 | 3.33 (2.95–3.71) |
| In-house | 24 | 64,931 | 2,009 | 3.63 (2.79–4.48) |
| **Race/ethnicity** |  |  |  |  |
| White, non-Hispanic | 7 | 114,544 | 5,662 | 3.76 (1.43–6.08) |
| Black, non-Hispanic | 7 | 7,287 | 649 | 9.96 (2.95–16.97) |
| Brown/Hispanic | 7 | 14,347 | 1,016 | 8.76 (0.01–18.65) |
| Multiple race/Asian/Other/Unknown | 7 | 8,139 | 709 | 5.78 (1.76–9.79) |

Of the 47 studies, 36 tested people of all age groups, whereas nine, and two studies tested only adults and children, respectively (Table 2). Subgroup analysis revealed pooled seroprevalences of 2.43% (2.16–2.70%), 5.31% (4.12–6.50%), and 8.76% (7.46–10.06%) in the general population, adults only and children only, respectively (Table 2).

### Seroprevalence in relation to serological assay used

Of 47 studies, 18 studies utilised rapid LFIAs to detect of specific serum antibodies against SARS-CoV-2, 11 used ELISA, 13 used CLIAs, four studies a virus neutralisation assay, and one used a microsphere immunoassay. Thirty-seven studies used commercial kits and 10 employed in-house serological methods. Subgroup analyses, conducted considering the type of serological method used, revealed pooled seroprevalences of 3.95% (3.17– 4.74%), 3.53% (2.65–4.40%), 2.73% (2.03–3.42%) and 1.32% (0.90–1.74%) using LFIA, ELISA, CLIA and neutralisation assay, respectively. One study in the USA, which used a microsphere immunoassay indicated a seroprevalence of 12.5% (11.97-13.03%). Subgroup analysis revealed pooled seroprevalences of 3.33% (2.95–3.71%) using commercial and 3.63% (2.79–4.48%) employing in-house assays (Table 2).

### Seroprevalence in relation to race

Seven studies (five from the USA, one from England and one from Brazil) had datasets that were stratified according to race. Subgroup analysis revealed pooled seroprevalences of 3.76% (1.43–6.08%); 9.96% (2.95–16.97%); 8.76% (0.01–18.65%); and 5.78% (1.76–9.79%) in white, black, Hispanic and other races (Asian/other), respectively (Table 2). For races in the USA, subgroup analysis revealed pooled seroprevalences of 4.11% (1.45–6.78%); 10.83% (4.81–16.85%); 12.79% (2.33–27.91%); and 5.86% (1.12–10.60%) in white, non-Hispanic; black, non-Hispanic; Hispanic; and other races (Asian/other), respectively.

### Relationship between seroprevalence and socio-demographic variables

Thirty-five studies represented countries with high income and very high HDI levels; 11 represented countries with upper-middle income levels and high HDIs; and one country had lower-middle income and medium HDI levels. There was no study from countries with low income and low HDI levels. Subgroup analysis (Table 3), according to income and HDI level, revealed higher seroprevalences in countries with high income (4.44%, 3.77–5.1%) and very high HDI levels (4.37%, 3.71–5.02%) than in countries with upper-middle income (1.31%, 1.02–1.59%) and high HDI levels (1.35%, 1.06–1.64%). Random-effects meta-regression analyses showed a significant, increasing trend in seroprevalence with increasing income (coefficient [*C*] = 3.10e-07; *P*-value = 0.09), and HDI (*C* = 0.131; *P*-value = 0.01) levels (Figure 3A-B).

**Table 3.** Prevalence of serum antibodies against SARS-CoV-2 in the general population based on sub-groups according to different socio-demographic and geographic parameters, calculated using a random effects model.

| Parameters/subgroups | Number of datasets | Number of people screened (total) | Number of seropositive people | Pooled seroprevalence % (95% CI) |
| --- | --- | --- | --- | --- |
| **Income** |  |  |  |  |
| Lower middle | 1 | 3,098 | 174 | 5.62 (4.83–6.49) |
| Upper middle | 26 | 120,242 | 2,361 | 1.31 (1.02–1.59) |
| High | 80 | 275,925 | 13,344 | 4.44 (3.77–5.1) |
| **Human Development Index** |  |  |  |  |
| Medium | 1 | 3,098 | 174 | 5.62 (4.83–6.49) |
| High | 26 | 119,426 | 2,358 | 1.35 (1.06–1.64) |
| Very high | 80 | 276,741 | 13,347 | 4.37 (3.71–5.02) |
| **Latitude** |  |  |  |  |
| 0-20° | 5 | 18, 007 | 496 | 2.99 (0.71–5.28) |
| 20-40° | 49 | 160,890 | 4,034 | 2.29 (2.03–2.56) |
| 40-60° | 53 | 220,368 | 11,349 | 4.68 (3.92–5.43) |
| **Longitude** |  |  |  |  |
| 0-30° | 53 | 220,851 | 9,969 | 4.15 (3.49–4.82) |
| 30-60° | 16 | 35,478 | 769 | 1.76 (1.18–2.34) |
| 60-90° | 10 | 27,927 | 2,435 | 6.36 (3.07–9.66) |
| 90-120° | 18 | 102,378 | 2,497 | 2.80 (2.37–3.22) |
| ≥ 120 | 10 | 12,631 | 209 | 1.63 (1.01–2.25) |
| **Relative humidity (%)** |  |  |  |  |
| < 60 | 15 | 62,692 | 3,763 | 5.84 (3.81–7.87) |
| 60-79 | 76 | 306,057 | 11,159 | 3.41 (2.96–3.85) |
| ≥ 80 | 16 | 30,516 | 957 | 2.77 (2.01–3.55) |
| **Mean temperature (°C)** |  |  |  |  |
| < 7 | 4 | 1,765 | 102 | 7.87 (1.54–14.20) |
| 7.1-13 | 36 | 111,683 | 5,351 | 4.27 (3.23–5.32) |
| 13.1-19 | 43 | 232,763 | 9,332 | 4.16 (3.53–4.78) |
| 19.1-25 | 18 | 29,550 | 303 | 0.85 (0.60–1.11) |
| 25.1-30 | 6 | 23,504 | 791 | 3.79 (1.75–5.84) |

**Figure 3.**
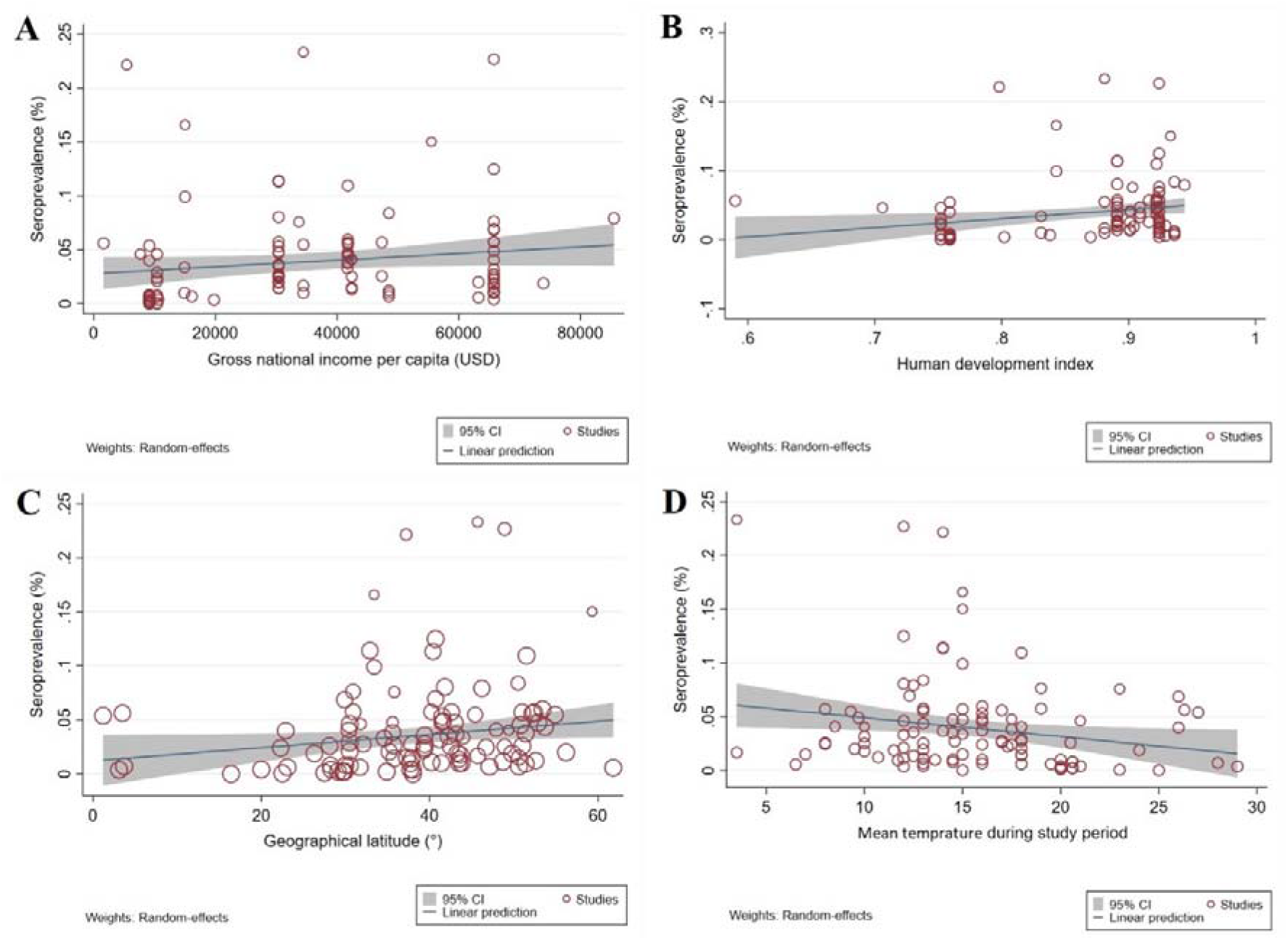
Ecological random effects meta-regression analyses of the seroprevalence of SARS-CoV-2 in the general population according to: (A) a country’s income level – showing a statistically significant upward trend in seroprevalence in countries with higher income levels; (B) human development index (HDI) – showing a statistically significant upward trend in seroprevalence in countries with higher HDIs; (C) geographical latitude – showing a statistically significant upward trend in seroprevalence with increasing geographical latitude; (D) the mean temperature during study implementation – showing a statistically significant downward trend in seroprevalence with increasing mean temperature

### Relationship between seroprevalence and geographical location or climate

At geographical latitudes of 0–20°, 20-40° and 40-60°, seroprevalences were 2.99% (0.71–5.28%), 2.29% (2.03–2.56%) and 4.68% (3.92–5.43%), respectively; the highest and lowest seroprevalences were at longitudes 60–90° (6.36%, 3.07–9.66%) and ≥120° (1.63%, 1.01–2.25%). In relation to climate, seroprevalences were 5.48% (3.81–7.87%), 3.41% (2.96– 3.85%) and 2.77% (2.01–3.55%) in regions with mean relative humidities of < 60%, 60-80%, and > 80%, respectively. Subgroup analysis indicated that the highest and lowest seroprevalences occurred in climes with mean environmental temperatures of < 7°C (7.87%, 1.54–14.20%) and 19-25°C (0.85%, 0.60–1.11%), respectively (Table 3). There was a significant (coefficient [*C*] = 0.0007; *P*-value = 0.03), increasing trend in seroprevalence with increasing geographical latitude (Figure 3C), and a non-significant (*C* = −0.00008; *P*-value = 0.316), decreasing trend with geographical longitude (Supplementary Figure 1A). Furthermore, there was a significant (*C* = −0.0017; *P*-value = 0.02), decreasing trend in seroprevalence with increasing, mean environmental temperature (Figure 3D) and a non-significant (*C* = −0.0006, *P*-value = 0.12), decreasing trend with increasing relative humidity (Supplementary Figure 1B).

### Association between seroprevalence and confirmed COVID-19 cases (i.e. disease) and death

Meta-regression analyses showed a non-significant, increasing trend in the number confirmed cases (*C* = 0.0002; *P*-value = 0.921) and mortality (*C* = 0.0001; *P*-value = 0.640) with increasing seroprevalences (Supplementary Figure 2A-B).

## Discussion

Currently, COVID-19 is the number-one public health concern worldwide. Here, we provide a comprehensive review of the seroprevalence of SARS-CoV-2 infection in general population from continents from which peer-reviewed investigations have been published at this time point (20 August 2020). The meta-analysis revealed a pooled seroprevalence of SARS-CoV-2 infection/exposure of 3.38% (95% CI, 3.05%–3.72%) relating to ~ 264 million individuals worldwide at the time of drafting this manuscript. Our findings are in accord with the World Health Organization (WHO) report projecting that 2-3% of the global population may have been infected by the end of the first epidemic wave [38]. Our findings also indicate that ~ 96% of the world’s population is still susceptible to COVID-19.

Overall seroprevalence varied markedly among countries and regions, which is attributable to many factors, including in cultural practices, political decision-making, policies, mitigation efforts, health infrastructure and prevention/control measures and/or the effectiveness of their implementation [39, 40]. Subgroup analysis showed higher seroprevalences rates in countries with higher income levels and HDIs. Due to lack of data for many disadvantaged countries, findings need to be interpreted with caution, but possible explanations might include increased urbanisation and population density, higher levels of social interaction and intensity of international travel. Morever, our analysis did not extend to a time when COVID-19 will accelerate in the Southern Hemisphere, especially in Africa and South America, or in South Asia. Therefore, suggested differences in the impact of COVID-19 due to differing national policies respect to Bacillus Calmette-Guérin (BCG) vaccination of children might not hold with this Southern expansion [39].

No significant gender or age difference in the seroprevalence of SARS-CoV-2 infection/exposure suggests that people of both sexes and all ages are at risk of infection, and that preventive measures should be implemented in the same way for all individuals. An interesting finding was the lower seroprevalence in white people compared with other racial and ethnic minority groups, which is in accordance with previous studies [41-44] reporting that minority groups are being disproportionately impacted by COVID-19. According to the Centers for Disease Control and Prevention (CDC) [45], factors contributing to this disproportionate impact include discrimination in health care, housing, education and finances; communication and language barriers; cultural differences between patients and health care providers; lack of health insurance; increased employment of racial and ethnic minority groups in essential work settings such as healthcare facilities, farms, factories, grocery stores, and public transportation; and living in more crowded family or conditions. In this analysis, we did not attempt to distinguish prevalence rates within different regions of the US or other nations. Doing so might have revealed increased seroprevalence in lower income areas due to the factors mentioned above. Indeed, it was noted that the “blue marble health” concept of poverty-related diseases amongst the poor living in high income nations might apply to COVID-19, just as it does for neglected tropical diseases, tuberculosis and other poverty-related conditions [46, 47].

Regarding the serological tests, our analysis indicated some variation in diagnostic sensitivity (detecting based IgG or total serum antibodies) among the serological assays used in published studies. A recent investigation indicated sensitivity and specificity of 85% and 99% for both IgG and total antibodies [48]. This review showed that similar seroprevalences were established by ELISA (3.9%) and LFIA (3.5%), while lower seroprevalence rates were obtained using CLIA (2.7%) and virus neutralisation (1.3%) assays. Two recent meta-analyses [49, 50] showed that sensitivities were consistently lower for the LFIA (66-80%) assay compared with ELISA (84-93%) and CLIA (90-97%), while a specificity of > 95% was calculated for all methods. Variation in sensitivity could be attributable to differences in the antigens used (i.e. recombinant or purified protein), the antibody conjugate employed and cut-off set for an assay [49, 51]. A Cochrane review indicated that the combination of the detection of IgG and IgM achieved a sensitivity of 30.1% one to 7 days, 72.2% for 8 to 14 days, 91.4% for 15 to 21 days after the onset of symptoms [52]. In present study [22, 25, 53, 54], four studies used virus neutralisation to detect serum antibodies to SARS-CoV-2, all exhibiting a sensitivity and specificity of > 98%. Neutralisation assays are more time consuming to perform (3–5 days) and are carried out in laboratories of a Biosafety Level-3 (BSL-3) [55]; therefore, these assays might be less suited for routine use. One study [56] used a microsphere immunoassay to detect serum antibodies to SARS-CoV-2 and indicated a high seroprevalence (12.5%). Although this method has been approved by the Food and Drug Administration (FDA), results might be interpreted with caution, given that only one study has been published to date.

With respect to geographical locations and climate parameters, we revealed an increasing trend for seroprevalence at higher latitudes and lower mean relative humidity and temperature. This finding is consistent with some previous laboratory, epidemiological and mathematical modelling studies [57-60], showing that environmental temperature and humidity play key roles in the survival and transmission of seasonal respiratory viruses. The highest seroprevalences were recorded at latitudes between 40°N and 56°N, in accordance with a previous study [60] showing substantial community spread of SARS-CoV-2 up to March 2020 in areas located in a narrow band in the 30° N and 50° N “corridor”. The finding of higher seroprevalence rates in areas with a low mean relative humidity and temperature accords with recent epidemiological [60] and laboratory [61] studies of coronavirus survival. Both environmental temperature and humidity are known as critical factors determining survival and community transmission of SARS-CoV, MERS-CoV and influenza [60, 62, 63]. Processes or mechanisms proposed to be linked to cold temperatures and low humidity include airborne droplet-stabilisation, increased viral replication in the nasopharyngeal mucosa or respiratory epithelium and/or reduced local innate immune responses, as evidenced for other respiratory viruses [57, 60, 64-66].

As the present study represents a first “snap shot” of seroprevalence based on a critical evaluation of published information, it has a number of limitations: First, one limitation is the lack of published, population-based studies from many countries across the globe at this relatively early phase of the pandemic, and some of the studies included here lacked data on sex and age of subjects tested. We hope that these limitations can be addressed over the next months and years, so that future estimates will be more representative of the situation worldwide, so that conclusions might be drawn regarding endemic stability and instability in particular countries and regions. Second, different serological methods/assays (with varying sensitivities and specificities) were employed in different studies, which will have some effect on the accuracy of our global estimate, although subgroup analyses were undertaken to assess a potential effect of the serological methods used. Third, pooled analyses showed significant heterogeneity. As such heterogeneity was expected in meta-analyses of global prevalence estimates [67-69], we explored possible sources of heterogeneity, including geographic region and diagnostic methods. However, we did not find the source of this heterogeneity.

This study reinforces that SARS-CoV-2 infection is a major global health threat and very rapidly-spreading communicable disease, with as the global seroprevalence rising to 3.38% only months after the commencement of the pandemic. Our findings suggest, though, that ~ 96% of the world’s population is still susceptible to infection, which is very alarming. This means that many countries could still face one or more surges in cases and, hence, overwhelm medical systems. We have seen in many locations that hospital beds, intensive care units (ICUs) and ventilators reached capacity. For example, early on, in New York city, the USA had to send mercy ships to handle the surge in need. Therefore, countries should have plans and medical resources in place for future, unexpected waves of COVID-19.

There are indications from many countries that mortality rates for COVID-19 are higher than those officially reported [70-72]. Hence, until a vaccine(s) is (are) available, the focus needs to be on education and prevention and strict quarantine measures. Indeed, universal masks and safe distance are our only means of reducing exposure, infections, disease and deaths. A global meta-analysis [73] showed that applying physical distancing of ≥ 1 m, usage of personal protective equipment (PPE, including a face mask, eye and body protection) results in a substantial reduction in the risk of transmission/infection. However, the lack of preparedness in many countries to control a rapid-spreading, high virulent and pathogenic virus, combined with limited or no biosecurity strategies/policies on how to deal with pandemics in populations, meant that such simple measures were not introduced initially. Our study calls for routine surveys to monitor temporal changes in seroprevalence in a location. In the context of epidemics and pandemics, such surveys might be conducted on a monthly or two-weekly basis to allow authorities to assess the spread of the virus and exposure levels in populations. A global plan is needed to monitor the seroprevalence of SARS-CoV-2 to assist prevention and control efforts. We aim to continue to follow the global seroprevalence situation over time, and to regularly report on trends and changes.

## Data Availability

All data related with this study are available in the manuscript and supplementary files.

## Contributors

A.R., R.B.G and P.J.H conceived the study. A.R., M.S., M.N.S. and S.E. conducted the searches and collected data. A.R., M.S., S.M.R. and M.L. analysed and interpreted the datasets. A.R., A.H.M, R.B.G and P.J.H drafted and edited the manuscript. All authors commented on, or edited drafts and approved the final version of the manuscript.

## Declaration of interests

All other authors declare no competing interests.

## Acknowledgements

AR’s research was supported by the Health Research Institute at the Babol University of Medical Sciences, Babol, Iran. RBG’s research is currently supported by the Australian Research Council (ARC), National Health and Medical Research Council (NHMRC) of Australia, Yourgene Health Singapore, and Melbourne Water. Thanks to Constantine E. Gasser for comments on the manuscript

## Notes

### Competing Interest Statement

The authors have declared no competing interest.

### Funding Statement

There is no funding source for this study

### Author Declarations

this study was approved by ethical committee of Babol University of Medical Science

